# PHARMWATCH: A Multilayer Pharmacogenomics Safety System for Accurate Star Allele Interpretation

**DOI:** 10.64898/2026.02.26.26347200

**Authors:** Chris Eisenhart, Rachel Brickey, Joel Mewton

## Abstract

The Clinical Pharmacogenetics Implementation Consortium (CPIC) bases its drug–gene recommendations on the assignment of star alleles, which map known genotypes to defined functional categories and corresponding drug dosage guidelines. The star allele framework, first proposed in 1996 for the *CYP* gene family and later formalized with CPIC’s establishment in 2010 [1, 2], remains foundational to pharmacogenomics. However, this system has notable limitations. Its dependence on a restricted set of benchmark single nucleotide polymorphisms (SNPs) excludes rare or novel pathogenic variants that can invalidate a star allele call and lead to incorrect dosing recommendations. Furthermore, nearby non-pathogenic variants can interfere with haplotype interpretation, introducing additional risk of misclassification.

To address these gaps, we developed PHARMWATCH, a multistep pharmacogenomics workflow for comprehensive variant analysis, allele tracking, and contextual interpretation. PHARMWATCH incorporates two algorithmic safeguards designed to identify genomic alterations that compromise star allele accuracy: (1) de novo germline variant screening using the ACMG-based BIAS-2015 classifier and (2) variant interpretation in context (VIIC) to validate the functional integrity of star allele–defining SNPs [3]. Together, these layers enhance the reliability of pharmacogenomic reporting, enabling safe, automated, and review-ready recommendations that extend beyond the constraints of traditional star allele–based approaches.

## 1 Introduction

In 2023, Dr. Anil Kapoor, internationally-renowned Canadian Professor of Urology and Head of Renal Transplantation at McMaster University, was diagnosed with colon cancer. In preparation for treatment with 5-fluorouracil, he underwent standard *DPYD* genotyping for the five most common variants, which returned negative—indicating no predicted risk of toxicity. Based on this result, he proceeded with 5-fluorouracil therapy. However, after a single dose, within days he suffered fatal toxicity, including severe gastrointestinal distress and multi-organ failure. Post-mortem next-generation sequencing (NGS) revealed a variant, *DPYD c.704G*>*A* that had previously been reported in the literature in another patient of South Asian ethnicity. This had been missed by the initial assay due to panel and technology limitations [4].

This case underscores the critical need for comprehensive NGS-based pharmacogenomic (PGx) testing that assesses all variants within key drug-metabolizing genes and evaluates their potential functional impact before guiding clinical decisions. While complete sequencing is essential for variant detection, standard evaluation of NGS data is insufficient. A robust interpretive framework is required to identify rare pathogenic variants and contextual interactions that may invalidate star allele–based classifications. Only through the combination of comprehensive sequencing and contextual interpretation could the *c.704G*>*A* variant have been recognized, potentially preventing Dr. Kapoor’s fatal outcome.

To address this unmet need, we developed **PHARMWATCH** (**P**harmacogenomics **H**andling, **A**nalysis, and **R**eporting with **M**ultistep **W**orkflow, **A**llele **T**racking, and **C**ontextual **H**armonization), a multilayer pipeline that enables accurate, automated, and review-ready pharmacogenomic recommendations consistent with CPIC guidelines. PHARMWATCH integrates three core components: CPIC’s PharmCat for standard star allele assignment, Bitscopic’s BIAS-2015 algorithm for germline variant classification under ACMG criteria, and Bitscopic’s Variant Interpretation in Context (VIIC) algorithm for verifying the biological integrity of star allele–defining variants [5].

At the core of the workflow, PharmCat processes variant call format (VCF) files using CPIC-derived tables [6]. The tool evaluates SNPs associated with each gene’s star allele definition, assigns corresponding alleles, and maps them to CPIC and DPWG drug dosage recommendations. However, PharmCat does not assess variants outside the predefined star allele framework, leaving the potential for missed pathogenic alterations or corruption of allele-defining sites by nearby genomic changes.

To close this gap, PHARMWATCH runs two algorithms in parallel with PharmCat; BIAS-2015 and VIIC. BIAS-2015 identifies any germline variants classified as pathogenic or likely pathogenic (P/LP) according to ACMG 2015 standards, flagging the affected gene for review. Concurrently, VIIC evaluates all variants contributing to star allele definitions and verifies that they produce the expected amino acid outcomes. This dual-layer validation ensures that star allele assignments unflagged by PHARMWATCH can be trusted with high confidence. By coupling variant-level pathogenicity screening with contextual interpretation, PHARMWATCH enhances the clinical reliability of pharmacogenomic testing and reduces the risk of preventable adverse drug reactions.

## 2 PHARMWATCH

### 2.1 *DPYD* in CPIC

The *DPYD* gene encodes dihydropyrimidine dehydrogenase (DPD), the rate-limiting enzyme responsible for metabolizing fluoropyrimidine chemotherapeutic agents such as 5-fluorouracil (5-FU) and capecitabine. Genetic variations in *DPYD* can markedly reduce DPD activity, substantially increasing the risk of severe or fatal toxicity in patients receiving these drugs [7]. The *DPYD* locus spans nearly one million base pairs on chromosome 1 (GRCh38/hg38: chr1:97,077,743–97,921,034), creating many opportunities for both inherited and de novo variants to occur [8].

Within the CPIC framework, 82 *DPYD* variants are currently incorporated into star allele definitions. By contrast, ClinVar reports more than 550 unique short variants for *DPYD*, meaning over 80% of ClinVar short variants are not evaluated in the *DPYD* star allele system. This can be seen clearly when examining exon 7 of *DPYD*. CPIC evaluates only a single SNP, rs1801266, for the classification of the star allele *\*8*. This rare variant produces the amino acid substitution p.R235W and has been associated with a complete reduction in enzyme activity. However, multiple additional variants have been reported in ClinVar for the same exon and are classified as pathogenic (Figure 1) [9]. These clinically significant alterations fall outside the star allele model and are therefore excluded from analysis, leading to potential mismatches between the assigned star allele function and the true enzymatic phenotype of the patient.

**Figure 1.**
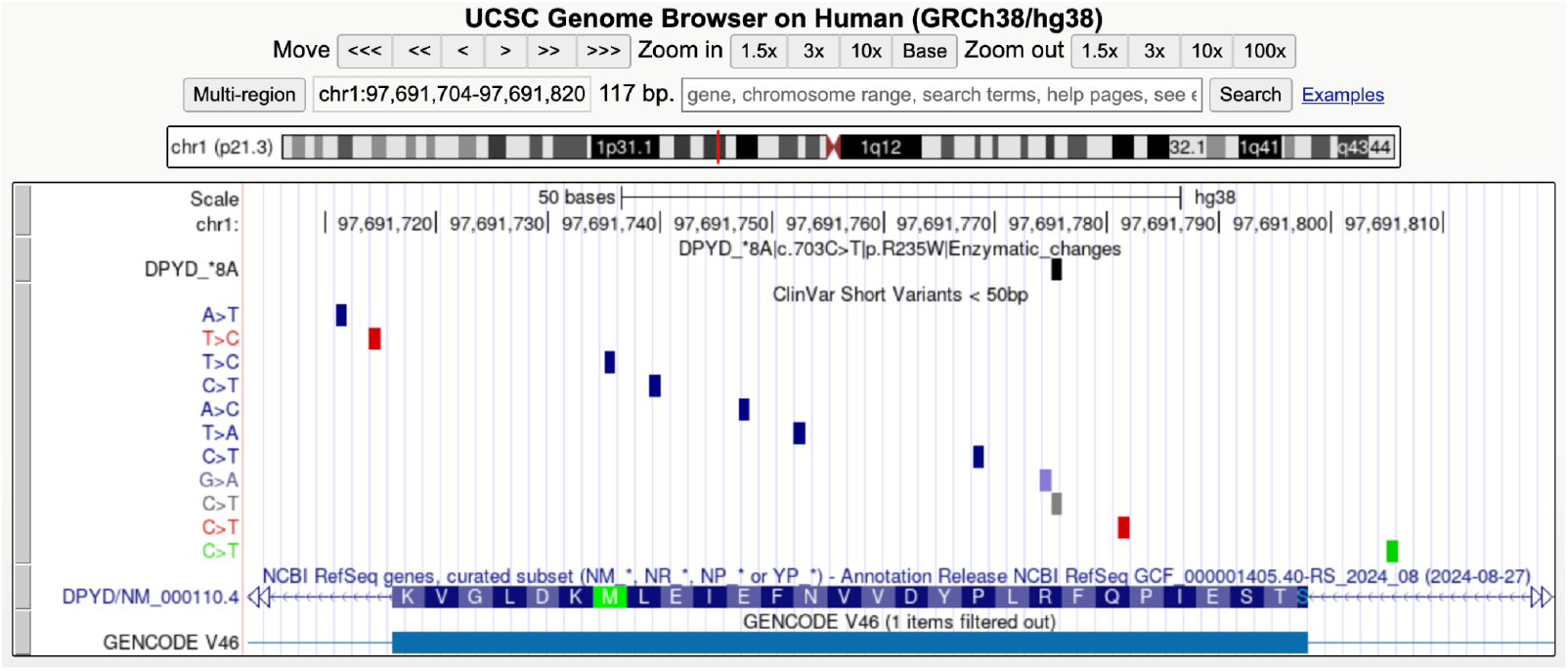
UCSC Genome Browser view of exon 7 (of 23) in *DPYD*. The top track shows the CPIC star allele–defining variant (rs1801266), the next track displays all variants reported in ClinVar (pathogenic variants in red), and the lower tracks depict exon structure and amino acid sequence from NCBI RefSeq [10].

### 2.2 Germline Variant Calling with BIAS-2015

BIAS-2015 (Bitscopic Interpreting ACMG 2015 Standards) is Bitscopic’s ACMG-based variant interpretation engine designed to automatically classify germline variants from next-generation sequencing (NGS) data [3]. The algorithm integrates multiple evidence streams, including population frequency data (gnomAD, TOPMed), functional prediction, evolutionary conservation, and established variant repositories (ClinVar, HGMD), to assign standardized ACMG/AMP 2015 pathogenicity codes. Each variant is evaluated across 19 automated ACMG criteria to produce a categorical interpretation: pathogenic, likely pathogenic, variant of uncertain significance (VUS), likely benign, or benign.

Within PHARMWATCH, BIAS-2015 functions as the first safety checkpoint, scanning the entire PGx gene for pathogenic or likely pathogenic germline alterations, independent of their presence in CPIC star-allele tables. When such variants are identified, PHARMWATCH flags the corresponding gene for review, superseding any downstream star-allele assignments that would otherwise be used to guide drug dosing.

In our analysis of the variants shown in Figure 2, BIAS-2015 correctly classified all known pathogenic *DPYD* variants—including *c.704G*>*A*—as pathogenic and flagged them for review. In contrast, a standard CPIC workflow limited to star-allele SNPs (e.g., rs1801266 defining *\*8*) would have only evaluated the rs1801266 genomic location, which is often wild type. This discrepancy illustrates the clinical risk inherent in relying solely on star-allele panels.

**Figure 2.**
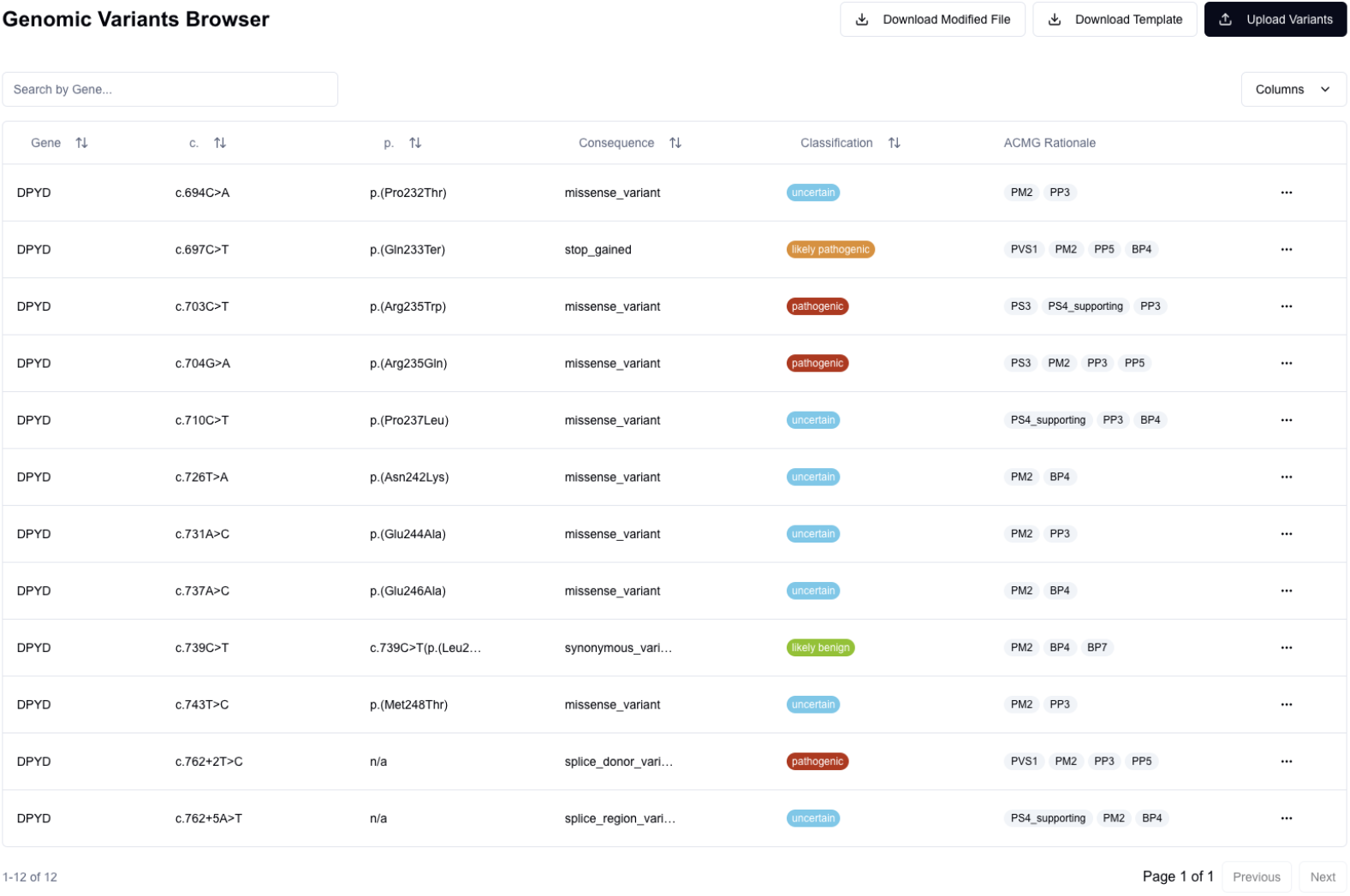
Twelve ClinVar variants covering exon 7 of *DPYD* displayed in the BIAS-2015 user interface following classification. The ACMG final interpretations (pathogenic, likely pathogenic, etc.) are shown alongside the supporting ACMG evidence codes. The *c.704G*>*A* variant is marked as pathogenic with PS3, PM4, PP3, and PP5 applied.

Importantly, BIAS-2015 can also evaluate de novo or previously unreported variants by applying predictive and contextual ACMG rules that do not rely on prior database annotation. Through conservation-based scoring, protein impact prediction, splice site analysis, and allele frequency modeling, the algorithm infers likely pathogenicity even for novel variants. This ensures that rare or uncharacterized alterations with potential functional consequences, such as a newly arising missense or splice variant in *DPYD*, are not overlooked. By extending interpretation beyond known cataloged variants, BIAS-2015 provides full-gene coverage and establishes a critical safety layer within pharmacogenomic workflows like PHARMWATCH.

Had BIAS-2015 been incorporated into Dr. Kapoor’s pre-treatment pharmacogenomic workflow, the *c.704G*>*A* variant would have been flagged for review before any CPIC-based functional assignment, prompting an immediate dose adjustment or consideration of an alternative therapy. This example underscores the clinical necessity of comprehensive germline screening prior to interpreting star-allele function: unrecognized pathogenic variants can invalidate a star allele classification even when NGS coverage over the relevant region is complete.

### 2.3 Variant Interpretation in Context (VIIC)

The Variant Interpretation in Context (VIIC) algorithm was developed to detect compound or contextual interactions between variants that can alter the expected functional outcome of a pharmacogenomic haplotype. Whereas tools such as BIAS-2015 evaluate each variant independently under ACMG guidelines, VIIC operates at the protein level—reconstructing and comparing amino acid sequences derived from the patient’s complete variant profile against those predicted from standard CPIC-defined star alleles.

For each pharmacogene region, VIIC generates three amino acid sequences: (1) the reference sequence from the reference genome, (2) the expected sequence predicted from the patient’s CPIC star allele assignments, and (3) the observed sequence derived directly from the patient’s NGS data. If the observed sequence deviates from the expected sequence at a star allele locus, the variant set fails VIIC validation, indicating a potential star allele corruption event that warrants manual review (Figure 3).

**Figure 3.**
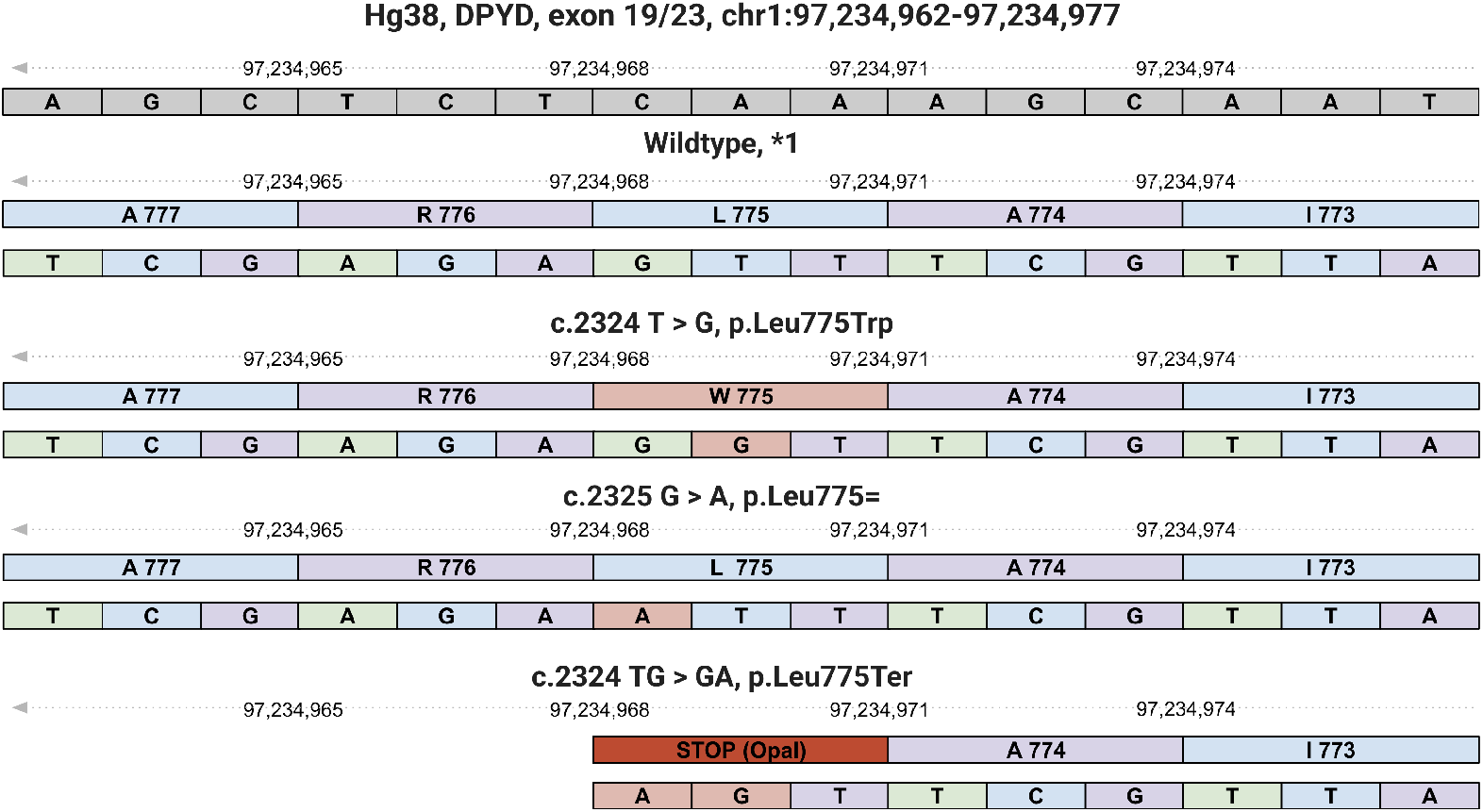
A section of exon 19 (of 23) in *DPYD* shown in standard genomic orientation. The gene is transcribed from right to left on the negative strand with reverse complement codons. Variant rs200643089 (*DPYD* p.Leu775Trp) alters the codon TTG (Leucine) to TGG (Tryptophan), producing a missense change. A nearby synonymous variant that changes TTG to TTA (Leucine → Leucine, p.Leu775=) would be benign in isolation. However, when both variants occur together, the codon becomes TGA, a premature stop codon, resulting in complete loss of function. This compound effect demonstrates how two individually benign or moderate variants can collectively disrupt protein function.

Such interactions would escape detection by BIAS-2015, since the synonymous variant alone would classify as benign and the CPIC star allele SNP would be excluded. Only through direct reconstruction and comparison of the encoded amino acid sequence does VIIC identify the emergent stop-gain effect. VIIC therefore complements BIAS-2015 by ensuring that amino acid changes resulting from multiple co-located variants are faithfully represented, preserving the biological validity of CPIC star allele assignments within PHARMWATCH.

Compound variant effects and multi-nucleotide variant (MNV) interactions have been increasingly recognized as a major challenge in clinical genomics, where nearby substitutions can collectively alter codon meaning or splicing outcomes [11, 12]. By explicitly reconstructing the true encoded sequence, VIIC operationalizes these emerging insights within a clinically deployable framework.

## 3 Methodology

The objective of the PHARMWATCH protocol is to ensure that every star allele definition applied to a gene can be validated with high confidence before being used to guide drug recommendations. To achieve this, PHARMWATCH introduces a two-step verification process that layers independent checks for variant pathogenicity and contextual sequence integrity, extending the reliability of standard CPIC workflows.

The protocol begins with PharmCat, which processes the variant call format (VCF) file to assign star alleles to each pharmacogene using CPIC-derived tables. Next, BIAS-2015 is executed on the same VCF file to classify all germline variants according to ACMG 2015 guidelines. If a pathogenic or likely pathogenic (P/LP) variant is detected within a gene that also has a defined star allele, and that variant is not part of the star allele’s definition, the corresponding star allele is considered corrupted and flagged for review.

Following this, the Variant Interpretation in Context (VIIC) algorithm evaluates all single nucleotide polymorphisms (SNPs) used in the star allele definition, including both variant and wild-type positions. If VIIC determines that any of these sites produce an unexpected amino acid outcome relative to the CPIC-defined sequence, the gene’s star allele is likewise flagged for review.

When PHARMWATCH is applied to a wild-type *DPYD* haplotype containing the c.704G>A variant, the gene is correctly flagged for review by both BIAS-2015 and VIIC contamination. When viewed through the PraediGene system, the biological mismatch between the assigned star-allele haplotype (predicting a wild-type Arginine) and the observed sequence (identifying a Glutamine substitution) becomes immediately apparent (Figure 4). This integration of automated contextual flagging and ‘human-in-the-loop’ review enables rapid turnaround times while ensuring rigorous safety checks for rare or novel genotypes that fall outside standard panels.

**Figure 4.**
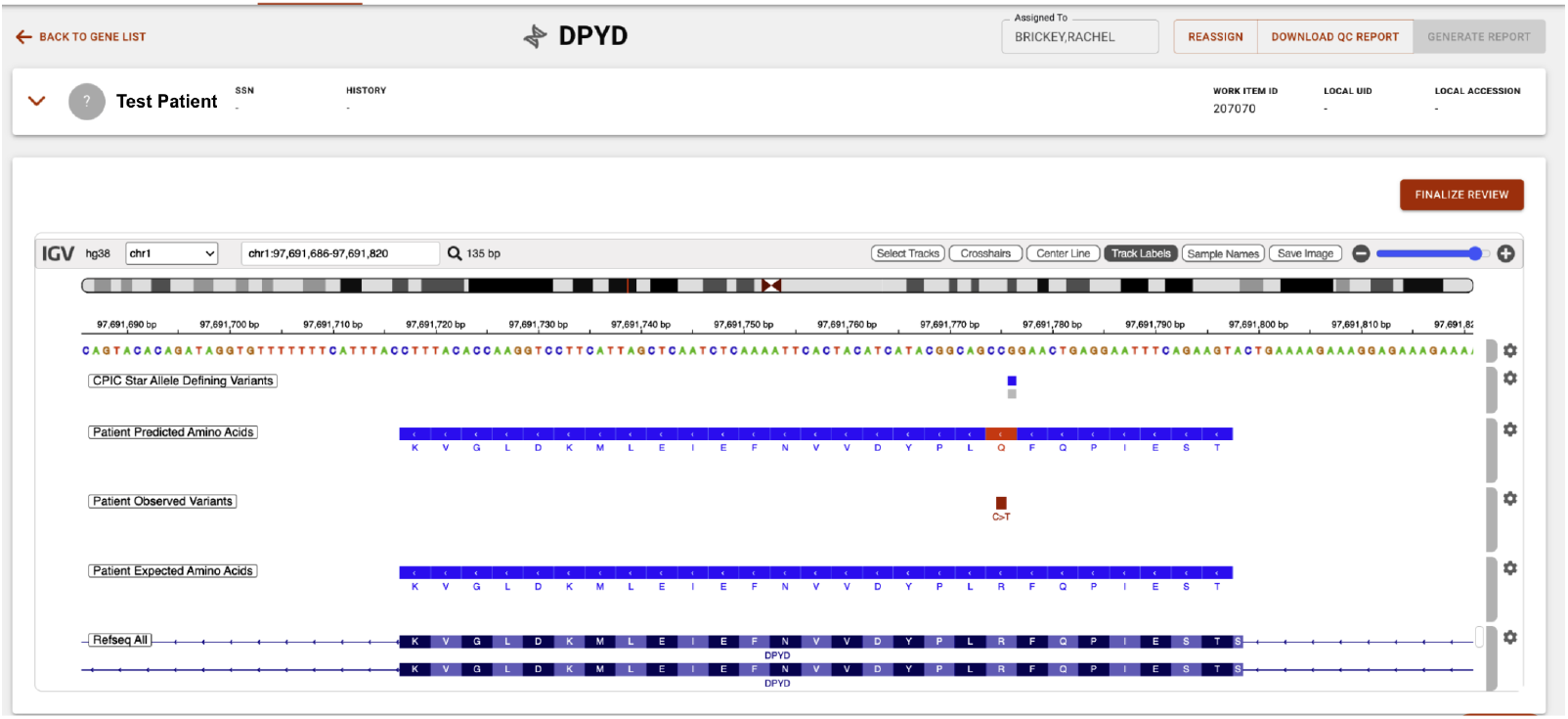
PraediGene VIIC Visualization of the *DPYD c.704G*>*A* Variant. The IGV-integrated browser displays the discordant amino acid reconstruction [13]. While the Patient Expected track (derived from standard CPIC star allele assignment) predicts an Arginine (R) at this locus, the Patient Predicted track (reconstructed from NGS data) identifies a Glutamine (Q) substitution. This automated flag enables the reviewer to identify a non-functional variant that would otherwise be masked by a wild-type star allele call.

Together, these overlapping checks establish two complementary validation layers. A single variant can be flagged by both algorithms—for instance, a pathogenic frameshift upstream of a star allele–defining SNP would be classified as pathogenic by BIAS-2015 and simultaneously identified by VIIC as corrupting the resulting amino acid sequence. Conversely, each algorithm can detect issues the other cannot: BIAS-2015 may flag pathogenic splice-site variants outside coding regions, whereas VIIC can identify compound interactions between otherwise benign variants that together alter protein function.

When a PHARMWATCH flag is raised by either BIAS-2015 or VIIC, the affected gene is annotated accordingly, and all relevant flags are included in the report for manual review. If no PHARMWATCH flags are detected, the PharmCat star allele assignments and corresponding CPIC/DPWG drug dosage recommendations are reported directly, indicating validation under the PHARMWATCH safety framework.

## 4 Limitations

### 4.1 General

PHARMWATCH is applicable only to pharmacogenes that use a star allele classification system with well-defined coding regions. With version 3.01 of PharmCat, this encompasses 34 genes. Additionally, PHARMWATCH applies only to genes whose star alleles are defined by single nucleotide polymorphisms (SNPs). Genes such as *HLA* or *CYP2D6*, in which star alleles are primarily determined by copy number variation, will not benefit from the PHARMWATCH workflow.

Furthermore, PHARMWATCH requires next-generation sequencing (NGS) data with broad and uniform coverage across the full gene locus. Microarray-based or targeted genotyping assays that measure only a predefined subset of SNPs do not provide sufficient variant context for PHARMWATCH to function correctly, since undetected variants cannot be evaluated for potential pathogenicity or contextual interference. Reliable interpretation therefore depends on comprehensive NGS data rather than limited SNP-panel inputs.

At present, PHARMWATCH flags anomalies for manual review. While effective, this process introduces additional turnaround time. Future versions aim to incorporate rule-based mapping of unique PHARMWATCH-flagged genotypes to their most probable functional outcomes and corresponding drug recommendations. This enhancement would allow PHARMWATCH to suggest or even resolve corrected star allele mappings automatically.

For example, consider a homozygous de novo loss-of-function (LOF) variant in *DPYD* within an otherwise wild-type background. Under the current CPIC framework, such a patient would be assigned a star allele of *\*1/*1*, indicating normal enzyme function and no dosage adjustment. PHARMWATCH, however, would flag the LOF variant for review. Because *DPYD* star alleles such as *\*2A* (c.1905+1G>A), *\*6* (c.1679T>G; p.Ile560Ser), *\*7* (c.299 302del; p.Val100fs), *\*12* (c.345C>A; p.Tyr115*), and *\*13* (c.1238G>A; p.Arg413Gln) all confer loss of function and recommend dose reduction for fluoropyrimidines, a future PHARMWATCH implementation could automatically map the novel LOF variant to an equivalent nonfunctional classification and propose an adjusted drug dosage recommendation.

### 4.2 BIAS-2015

BIAS-2015 v2.1.1 applies ACMG 2015 guidelines to classify variants into five standard categories: pathogenic (P), likely pathogenic (L/P), variant of uncertain significance (VUS), likely benign (L/B), and benign (B). As a strict implementation of the ACMG criteria, BIAS-2015 mirrors both the strengths and limitations of the framework: if the ACMG rules fail to capture a variant’s clinical context, BIAS-2015 will also be constrained. This differs from non-ACMG heuristic classifiers such as Sherloc or PrimateAI, which directly infer pathogenicity through empirical modeling [14, 15].

Benchmarking of BIAS-2015 v2.1.1 demonstrated sensitivity and specificity of 73% and 88% for pathogenic variants, 80% and 96% for benign variants, and 72% and 76% for uncertain variants, respectively. A detailed comparison with InterVar and full benchmarking results are provided in the BIAS-2015 manuscript [16].

### 4.3 VIIC

The VIIC algorithm relies on the UCSC Consensus Coding Sequence (CCDS) track for HG38 to define the canonical amino acid sequence for each evaluated genomic region [17]. Consequently, its analysis is limited to the canonical transcript represented in the CCDS dataset.

Although most variant callers can merge nearby variants based on gap thresholds and phasing parameters, this behavior only holds when variants share identical allele frequencies and read support. If one variant is homozygous and the other heterozygous, variant callers will report them as separate events, even when they occur within a single base pair.

This issue extends beyond zygosity differences. Many variant callers require all supporting FASTQ reads to fully overlap and agree for a multi-nucleotide variant (MNV) to be reported as a single entity. Even minor discrepancies among supporting reads can prevent proper merging. VIIC overcomes this limitation by jointly evaluating all proximal variants during translation reconstruction, ensuring that the combined amino acid consequences are accurately represented even when standard variant callers fail to merge the events.

## 5 Conclusion

PHARMWATCH addresses critical limitations of the CPIC star-allele framework, which depends exclusively on a narrow set of benchmark SNPs for haplotype assignment. In *DPYD*, the key gene governing fluoropyrimidine (5-FU) toxicity, this approach frequently overlooks rare pathogenic variants outside the core set (*\*2A*, HapB3, *\*9B, *13*) and fails to account for contextual interference from nearby variants that can distort star allele interpretation. For instance, exon 7 contains a single star-defining variant (*c.703C*>*T, *8*) but multiple additional pathogenic variants (e.g., *c.704G*>*A*) that standard pipelines exclude from evaluation.

PHARMWATCH mitigates these gaps by integrating two independent validation layers: BIAS-2015, which flags ACMG-classified pathogenic or likely pathogenic germline variants across the entire gene, and VIIC, which verifies amino acid outcomes for each star-defining SNP to ensure haplotype integrity. Together, these complementary algorithms safeguard against both missed pathogenic variants and star allele corruption due to contextual interference.

Had PHARMWATCH been applied to Dr. Anil Kapoor’s pre-treatment NGS data, the fatal *DPYD c.704G*>*A* variant would have been flagged prior to dosing, prompting review and dose adjustment. By combining comprehensive germline screening with contextual sequence validation, PHARMWATCH transforms pharmacogenomic testing from a limited tag-SNP screen into a multilayered safety frame-work—enabling accurate, automated, and clinically reliable CPIC-guided prescribing at scale.

## Declarations

### Ethics Statement

This study utilized publicly available genomic data (ClinVar, gnomAD) and previously published clinical reports. No original human subject research was conducted by the authors; therefore, Institutional Review Board (IRB) approval was not required. Regarding the clinical case description, informed consent for the inclusion of these details was provided by the subject’s next of kin.

### Data Availability

Genomic variant data used in this analysis were obtained from publicly available repositories, including ClinVar and the UCSC Genome Browser. The BIAS-2015 algorithm is available as an open-source tool. The PHARMWATCH workflow and VIIC algorithm are proprietary developments of Bitscopic, Inc.

## Acknowledgments

The authors would like to thank the family of Dr. Anil Kapoor for their assistance in providing clinical context and clarifying the details of his case.

